# Alternative protocols of tissue fixation dramatically reduces the impact of DNA artifacts, unraveling the interpretation of clinical Comprehensive Genomic Profiling

**DOI:** 10.1101/2023.06.17.23291533

**Authors:** Enrico Berrino, Sara Erika Bellomo, Anita Chesta, Paolo Detillo, Albero Bragoni, Amedeo Gagliardi, Alessio Naccarati, Matteo Cereda, Gianluca Witel, Anna Sapino, Benedetta Bussolati, Gianni Bussolati, Caterina Marchiò

**Affiliations:** Department of Medical Sciences, University of Turin; Candiolo Cancer Institute, FPO-IRCCS, Candiolo, TO, Italy; ADDAX Biosciences srl., Turin, Italy; IIGM-Italian Institute for Genomic Medicine, c/o IRCCS, Candiolo, TO, Italy; Department of Molecular Biotechnology and Health Sciences, University of Turin, Turin, Italy

**Keywords:** formalin fixation, sequencing, targeted panels, DNA integrity, precision medicine.

## Abstract

Formalin-fixed-paraffin-embedded (FFPE) samples represent the cornerstone of tissue-based analysis in precision medicine. Targeted next-generation sequencing (NGS) panels are routinely used to analyze a limited number of genes to guide treatment decision making for advanced stage patients. The number and complexity of genetic alterations to be investigated are rapidly growing, in several instances a comprehensive genomic profiling (CGP) analysis is required. The poor quality of genetic material extracted from FFPE samples may impact the feasibility/reliability of sequencing data.

We sampled 9 colorectal cancers to allow 4 parallel fixations: i) neutral buffered formalin (NBF); ii) acid-deprived formalin fixation (ADF); iii) pre-cooled ADF (*cold*ADF); iv) Glyoxal Acid-Free (GAF). DNA extraction, fragmentation analysis and sequencing by using two large NGS panels (OCAv3, TSO500) followed. We comprehensively analyzed library and sequencing QCs, and the quality of sequencing results.

Libraries from *cold*ADF samples showed significantly longer reads than the others with both panels. ADF and *cold*ADF derived libraries showed the lowest level of noise and the highest levels of uniformity with the OCAv3 panel, followed by GAF and NBF samples. The data uniformity was confirmed by TSO500 results, which also highlighted the best performance in terms of total region sequenced for ADF and *cold*ADF samples. NBF samples had a significantly smaller region sequenced, displayed a significantly lower number of evaluable MS loci and a significant increase in SNVs compared to other protocols. The mutational Signature 1 (aging-, FFPE artifact-related) showed the highest (37%) and the lowest (17%) values in NBF and *cold*ADF samples, respectively.

Most of identified genetic alterations were shared by all samples in each lesion. Five genes showed a different mutational status across samples and/or panels: four discordant results involved NBF samples.

In conclusion, acid-deprived fixatives guarantee the highest DNA preservation/sequencing performance, suggesting possible implementation of even more complex molecular profiling on tissue samples.

## INTRODUCTION

Next-Generation Sequencing (NGS) technology allows the detection and assessment of genetic alterations in cancer tissues, and it represents an increasingly practiced approach to meet the requirements of tailored therapies. Even though DNA from fresh-frozen tissues would appear as ideally suited for this type of analysis, several reasons impose the use of DNA extracted from routinary formalin-fixed paraffin embedded (FFPE) tissue blocks^1^. Notable is the bonus of the availability of well characterized archival material, which is collected with standard, reliable and time-honored procedures, and even stored for long times (up to several years). In addition, histological and, in case, immunohistochemical control allows to select and accordingly extract the correct tumoral lesion. Finally, a FFPE tissue block is often the only material available for properly addressing therapy-related questions.

Unfortunately, the poor quality of genetic material extracted from FFPE tissue blocks may impact the feasibility and the reliability of NGS data ^2–4^. Among the different pre-analytical steps involved in the generation of FFPE tissue blocks, the most critical one is represented by formalin fixation. Inadequate procedures, such as delayed, insufficient, or over-extended fixation or the use of acidic formalin are known to impact the results^5^. However, even following the standard use of Neutral Buffered Formalin (NBF), extensive DNA fragmentation and chemically induced changes are often affecting the results.

Formaldehyde binds to the amino groups of nucleotide bases. Deamination of cytosine can lead to misinterpretation of DNA sequences, particularly to an increased identification of cytosine (C) to thymine (T) and guanine (G) to adenine (A) (C:G > T:A). Several studies have investigated the detection of artifactual mutations resulting from the deamination process, a potential cause of erroneous treatments^1–4^. However, false-positive mutations remain a risk for NGS analysis of DNA extracted from FFPE tissue blocks, further enhanced by the extensive fragmentation of this material^3^.

Indeed, tissue fixation in formalin is known to produce DNA fragmentation, but the degree of fragmentation varies in different tissue blocks and, in this respect, the DNA “quality” (i.e., the degree of fragmentation) heavily impacts the results of genetic analysis. Several studies indicate that a reduced size of DNA templates decreases the success rate of amplicon-based methods and even leads to false positive data ^6–8^.

The formulation of NBF consists of a solution in phosphate buffer of commercial formaldehyde, a reagent known to be rich in formic acid. The latter, once linked to sodium ions in NBF, is commonly regarded as ineffective. However, we have recently shown that removal of formic acid is responsible for a significant improvement in DNA preservation^9^, pointing out possible detrimental effect of buffered acid residues on DNA, This was also supported by the evidence that a tissue fixation based on an acid deprived glyoxal, which is still a di-aldehyde, can be approached by maintaining the histological and immunophenotypical features of tissues^10^. Since there is a gap of knowledge on the real effect of tissue fixation on sequencing results, especially when sequencing is approached by means of a comprehensive genomic profiling (CGP) rather than by small, targeted panels, we have addressed this issue in the present study.

## MATERIALS AND METHODS

### Cases

We sampled 9 colorectal cancer patients with surgical resection of at least 2 cm in size to allow 4 parallel fixations (in total 36 FFPE tissue blocks). We also sampled a liver hepatocellular carcinoma (aka hepatoma) that underwent thermal-ablation before surgery, as a control of potential “necrosis-induced” DNA degradation.

The study was approved by the Ethics Institutional Review Board (IRB) responsible for “Biobanking and use of human tissues for experimental studies” – Department of Medical Sciences, University of Turin. Surgical specimens were processed following the under-vacuum sealing and cooling procedure^11, 12^. Each of the 10 collected samples was fixed for 24 hours in parallel as follows: (i) standard formalin fixation, i.e., NBF (Diapath, Bergamo Italy), pH 7.2-7.4, which represents the fixative used in daily practice and in all the previous projects^9, 10, 13^; (ii) acid-deprived formalin fixation, i.e., ADF, prepared by Addax Biosciences srl, (Turin, Italy) as previously reported^9^; (iii) pre-cooled ADF i.e., *cold*ADF, in which the sample was immersed in pre-cooled 4% ADF and fixed for 24h at 4°C, as previously described^13^; (iv) Glyoxal Acid-Free, i.e. GAF, prepared by Addax Biosciences s.r.l. (Turin, Italy), as previously described^10, 14^, for a total of 40 samples. Features of the fixatives were reported in Table 1.

**Table 1:**
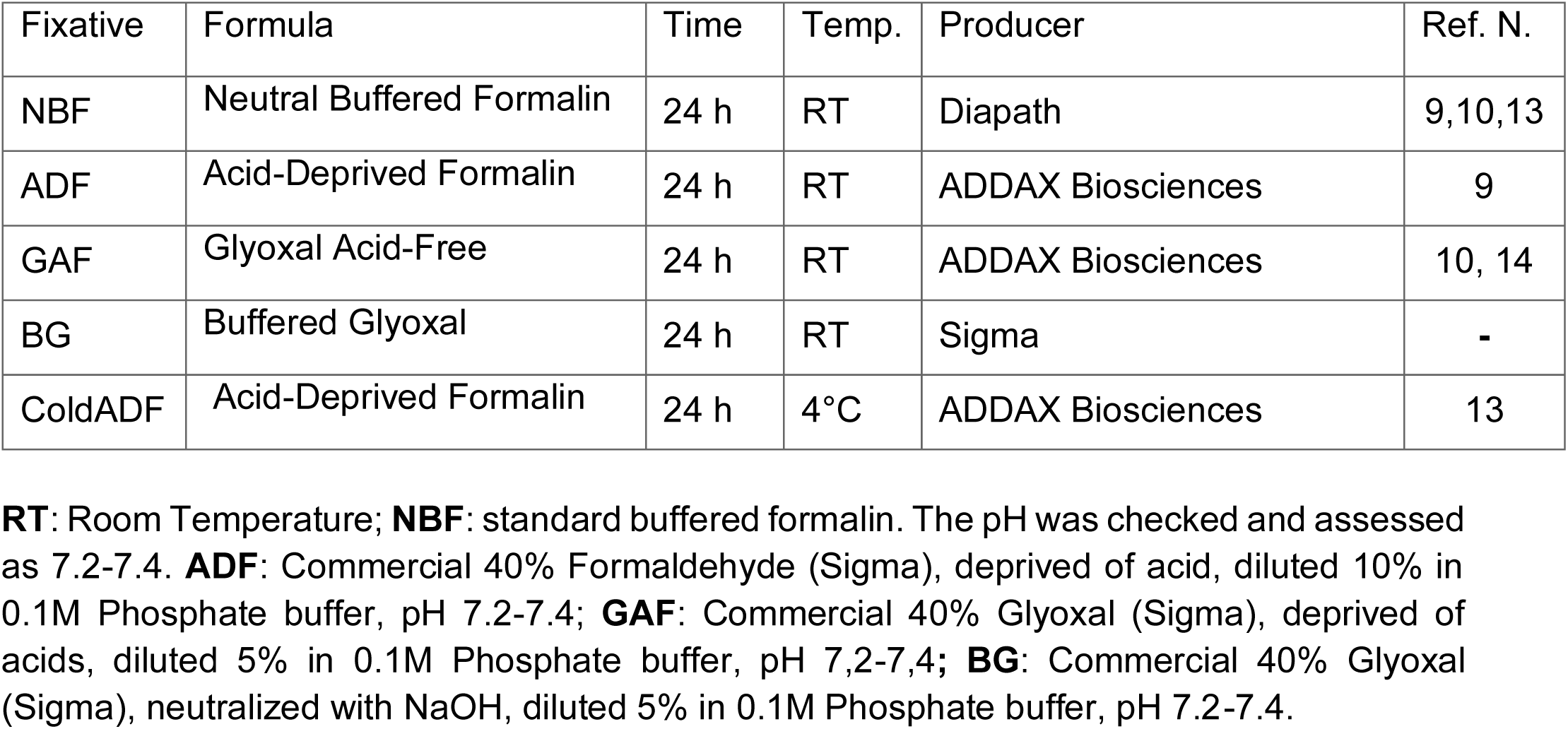
Reagents and fixation protocols.

A second cohort of nine tissues (both normal and tumoral specimens) were fixed only with GAF and with “standard glyoxal” (a solution of commercial Glyoxal in 0,1M Phosphate buffer pH 7,2) for a total of 18 tissue blocks.

After fixation, the samples were routinely processed and embedded in paraffin using an HistoCore PELORIS 3 Premium Tissue Processing System (Leica Biosystems, Italy)

### DNA Extraction and Quantification

Two pathologists evaluated histological and pathological features of tissue samples. Based on the hematoxylin and eosin (H&E) stained slide, we recorded two parameters: (i) the percentage of tumor area (%) and (ii) the total (mm^2^) tumor area. DNA was purified from five 6-µm thick sections with QIAamp DNA FFPE Advanced Kit (Qiagen, Hilden, Germany), following manufacturer’s protocol, and eluted in 40µL of nuclease-free water. We performed both fluorimetric (Qubit 3.0 fluorimeter, ThermoFisher Scientific, Wilmington, DE, USA) and spectrophotometric (NanoDrop 1000 Spectrophotometer ThermoFisher Scientific, Wilmington, DE, USA). The total yield of the extraction (ng) was normalized for the lesion area (mm2), and the ng/mm^2^ was used as the variable to define the extraction yield. Absorbance ratio between 260 nm and 230 nm (R230) and 280 nm (R280) were calculated by spectrophotometer to evaluate the extraction purity.

### DNA Fragmentation assays

As part of the pre-analytical Quality Controls (QCs) we assessed the DNA fragmentation with two independent methods.

On one side, we applied the Genomic DNA ScreenTape assay on the Agilent TapeStation 4150 instrument (Agilent Technologies, Santa Clara, CA, USA) allowing fragment analysis for a range between 0–60.000 bp for analysis. This method provides a discrete parameter for the DNA integrity (the DNA Integrity Number/DIN) and a continuous distribution of the fragments. The continuous size distribution analysis was performed as described here (^9^), calculating the total area under the curve for each bin of size.

On the other side, we applied the DEPArray™ FFPE QC Kit (Menarini-Silicon Biosystem, Bologna, Italy), as previously reported^13^. Briefly, the assay is based on two qPCRs, encompassing the same genomic region, but producing amplicons of different size (54 and 132 bp). Standard curves allow quantifying the amount of each primer, and the ratio between these amounts returns a linear QC score, ranging from 0 to ∞, where zero represents a highly fragmented DNA. The assay also represents a proof of amplifiability of the DNA sample.

### CGP by amplicon-based targeted sequencing

One hundred ng of each sample was used as input for the Oncomine Comprehensive Assay (OCAv3) targeted gene panel. This kit evaluates hot spot regions of 87 and the full coding region of 48 genes (gene for the DNA assay 135 genes), for a total of 0.397 Mb. The libraries were prepared using the Ion AmpliSeq Library kit Plus (Thermo Fisher Scientific) with the standard protocol. Samples were barcoded using 40 different Ion Xpress Barcodes. The Ion Library TaqMan Quantitation Kit (Thermo Fisher Scientific) allows a sensible and specific quantification to dilute each library to 100 pM. Samples were loaded into two 550 Ion Chip GeneStudio S5 Plus System for the sequencing (Thermo Fisher Scientific) with a minimum mean read depth of 500*x*. Aligned files (BAM) were processed from raw data and generated by the Ion Torrent software pipeline (Torrent Suite Software 5.12, Thermo Fisher Scientific). QCs and DNA variant calls were processed using the Ion Reporter Software (version. 5.10) and analyzed as previously reported^15^.

We assessed the quality of the sequencing by analyzing the following parameters:

1. Library QCs:

- Library yield. The quantity of library obtained by the on Library TaqMan Quantitation Kit (Thermo Fisher Scientific), in nM;
- Library size. The median size calculated using the DNA1000 HS of the Agilent TapeStation 4150 instrument (Agilent Technologies, Santa Clara, CA, USA), in bp;
2. Sequencing QCs:

- Read depth: the median value of sequencing depth, in *x*;
- Read length: the mean length of the insert size post reads trimming, in bp;
- Reads on target: the percentage of the reads aligned to the targeted regions, in %;
- Uniformity of coverage: the percentage of the analyzed genome in which the read depth is greater than 0.2 times the mean depth, in %;
- Median Absolute Pairwise Difference (MAPD). The median of the absolute values of all such differences in log_2_(*read count ratio*) is the measure of sequencing noise, important for cnv analysis. Higher values = reduced quality.
3. Sequencing results

- OncoPrint of Variants: representation of variant calling for all somatic SNVs and small indels with a variant allele frequency (VAF) >= 0.1.

### CGP by hybrid captured-based targeted sequencing

We performed DNA sequencing on the same samples by using a second NGS targeted panel, the Illumina TruSight Oncology 500. This hybrid captured-based approach comprises the coding sequence of 523 genes, with a total panel size of 1.94 Mb, 1.2 of coding regions. The size of this panel allows to evaluate tumor mutational burden and microsatellite instability, by processing the raw data using the associated Illumina local app. Briefly, 80 ng of genomic DNA was used to generate libraries that were sequenced on the Novaseq6000 instrument (Illumina, San Diego, California, USA) to reach a minimum of 150× read depth. Data was processed as previously described^16–18^. TSO500 data also allows to predict the mutational signatures for each type of fixation, by applying the MuSiCa tool for targeted sequencing ^19^. We assessed the quality of the sequencing by analyzing the following parameters:

1. Sequencing QC:

- Read depth: the median value of sequencing depth, in *x*;
- Read length: the mean length of the insert size post reads trimming, in bp;
- Coding region sequenced (Mb): the size of coding region sequenced with at least 50X of depth, in Mb;
- Usable MSI sites: the number of high-quality microsatellite sites;
- Chimerism: the variability of the libraries on the basis of the number of amplicon family and the percentage of chimeric reads, demultiplexing the unique molecular identifier (UMI) inserted during the library preparation;
- Reads on target: the percentage of the reads aligned to the targeted regions, in %;
- Uniformity of coverage: the percentage of the sequenced region covered at 100*x* and 250*x*, in %.
2. Sequencing results:

- Jaccard index (JI): a measurement of the similarity between the nucleotide alteration compared to the reference sequence in all the chromosome location, as previously described ^20^. We calculated the JI by comparing each fixation type on the basis of all nucleotide variants, both in germline and somatic settings;
- Mutational signature profile: the mutational signatures were evaluated using the six substitution subtypes (C>A, C>G, C>T, T>A, T>C, and T>G) with a VAF > 0.05 and their and their neighboring sequences ^21^;
- ncoPrint of variants: representation of variant calling for all somatic SNVs and small indels with a variant allele frequency (VAF) >= 0.1.

### Statistical analysis

Statistics were performed with the R software v4.03. Differences in distributions were analyzed with a paired T-test and contingency by Fisher’s exact or χ-square test. P < 0.05 were considered as statistically significant. The JI was calculated with the proxy (version 0.4-16) package of the R software.

## RESULTS

### Impact of the different fixations on DNA pre-analytic features

We conducted a series of tests on DNA from 9 colorectal carcinomas in order to compare the effect of fixation with standard NBF and two acid-deprived fixatives (ADF and GAF). In addition, we tested whether ADF pre-cooling could further ameliorate DNA preservation. We successfully purified DNA from all tissue samples (n=40). First, we assessed whether the different protocols could affect the yield of extraction. To reduce the impact of the lesion size, we normalized the total DNA extracted for the size, obtaining a value representing ng/mm^2^ for both fluorometric and spectrophotometric assays. Median levels were comparable for all the fixation types (paired T-test, Supplementary Figure 1A). In line with the DNA extraction yield, the different fixation types did not impact the R230 and R280 values (Supplementary Table 1, Supplementary Figure 1B).

To analyze in depth the degree of DNA fragmentation, we applied two parallel tests. The qPCR-based assay returned a QC score that was higher for ADF, *cold*ADF and GAF compared to NBF specimens, significantly higher in the comparison ADF *versus* NBF and *cold*ADF *versus* NBF (Supplementary Table 1 and Supplementary Figure 1C). The automated electrophoresis Tapestation 4150 provides both a punctual (DIN) and a continuous information about the fragment size distribution within the sample. *Cold*ADF showed the highest DIN values overall, even though differences among the different fixatives were not statistically significant (Supplementary Table 1 and Supplementary Figure 1C).

Since the DIN represents a static parameter, to comprehensively evaluate the composition of DNA fragments within the samples, we applied unsupervised clustering to the AUC of each bin size (Figure 1). We identified three classes of fragmentation, mostly related to the percentage of fragments < 1000 bp, with a trend of lower fragmentation from the top to the bottom of the heatmap (Figure 1). By annotating for the fixation type we identified a polarization of ADF and *coldADF* in the class with lower fragmentation (red and black squares in the left annotation of Figure 1). This was confirmed by χ-square test (p = 0.0009). This polarization was patient-independent, with the exclusion of DNAs purified from the thermo-ablated hepatoma, characterized by the lowest DINs and QC scores (right annotations in Figure 1) and clustering all together at the top of the heatmap (Figure 1). Taken together, these results suggest an increment in terms of purity and structural integrity of the DNA after acid deprivation (from NBF to ADF/*cold*ADF). We wondered whether this could be applied also to tissues fixed in standard glyoxal versus GAF (acid deprived glyoxal). Boxplots in Supplementary Figure 1D showed a significantly higher DIN in GAF specimens compared to those fixed with standard glyoxal.

**Figure 1.**
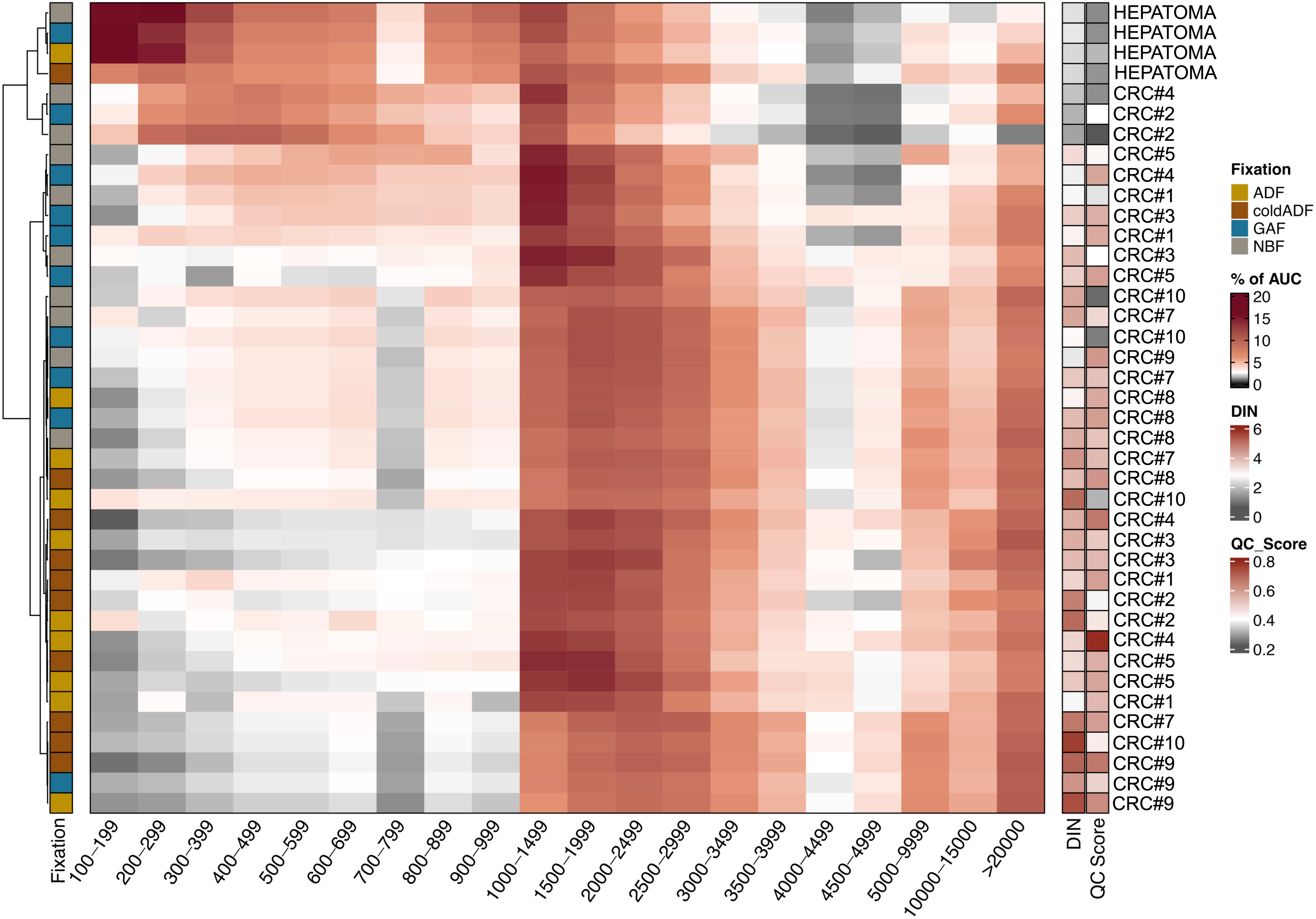
Heatmap of fragmentation spectra from TapeStation data. Each raw represents a single sample, with DNA size bins in columns (in bp). The reported parameter is the relative contribution (%) of each bin within the samples. Samples are sorted by unsupervised clustering, annotated on the left-hand side for the fixation type and annotated on the right-hand side for the DIN and the QC Score. AUC: area under the curve, DIN: DNA integrity number.

### Impact of the different fixations over a CGP based on an amplicon-based DNA targeted sequencing

Comparative results of the sequencing with the Oncomine Comprehensive v3 panel were divided into library preparation QCs, sequencing QCs and sequencing results.

In terms of library QCs, the analysis on the TapeStation electrophoresis showed larger library size (bp) for *cold*ADF-fixed specimens, confirming higher quality of these samples, despite a similar library yield (Supplementary Figure 2A). Boxplots displayed comparable library size for GAF and ADF, slightly higher than NBF-fixed samples.

We pooled the normalized libraries in two IonTorrent 550 chips, to reach at least a mean depth of 500x. Libraries were balanced, hence no differences in depth were identified across the different fixation types (Supplementary Table 2). After trimming, we detected a read length in line with the previous result of library size, with NBF-derived libraries significantly shorter than the others (Figure 2A, Supplementary Table 2). Following alignment, we calculated the fraction of reads falling on the target region, together with the uniformity of spreading within that region. Despite a comparable, high level of on-target sequencing (Figure 2B, Supplementary Table 2), uniformity was significantly reduced in NBF fixed tissues. Conversely, ADF and *cold*ADF derived libraries were characterized by high levels of uniformity, while GAF libraries by an intermediate level (Figure 2B, Supplementary Table 2).

**Figure 2.**
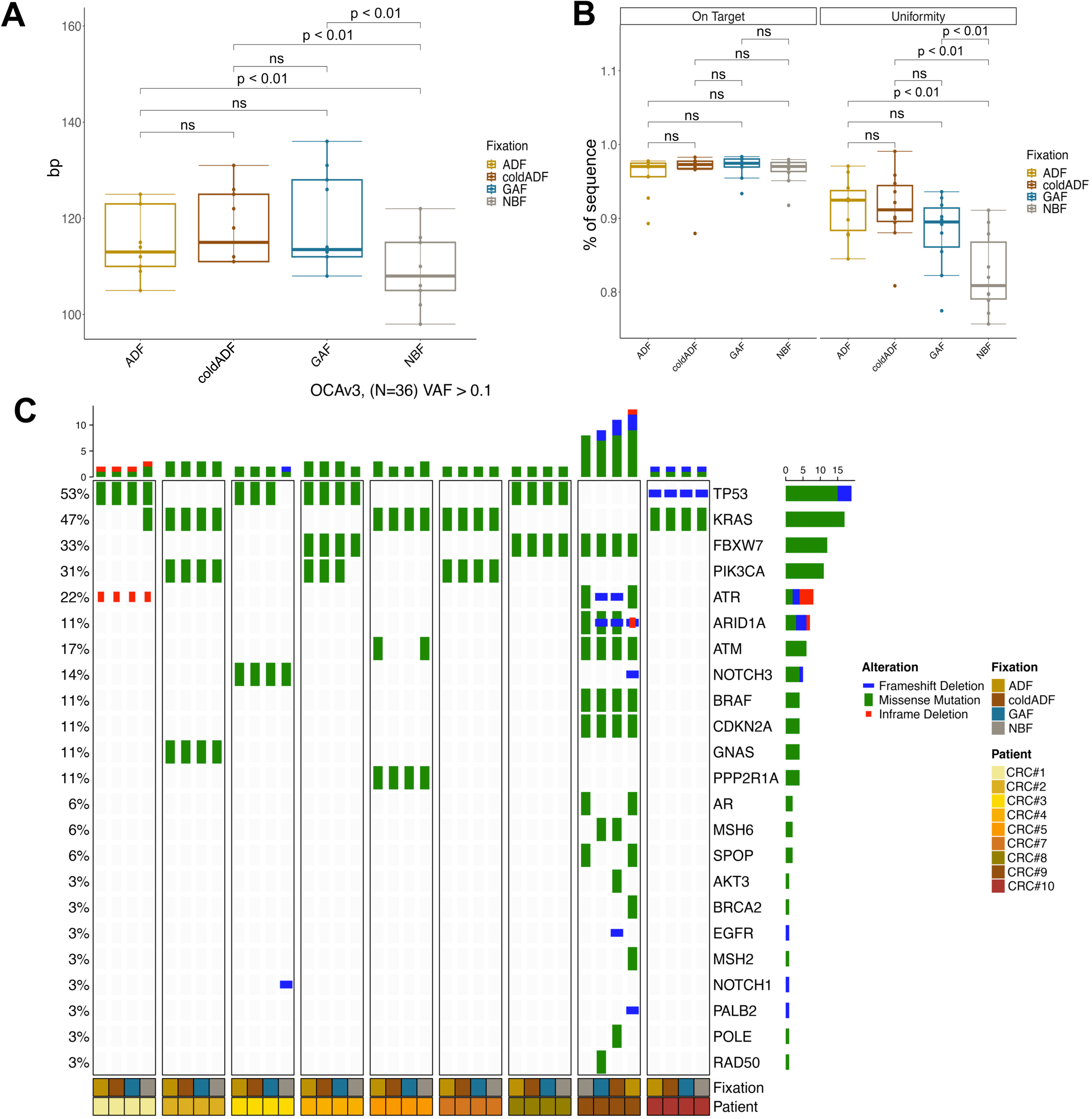
Output data by the OCAV3 panel. A) Boxplot of the OCAV3 read insert size. Y-axis reports the mean size for each sample, grouped by the fixation protocol. P-value is calculated with paired T-test. **B) Boxplots of the percentage of on target sequence and the uniformity of coverage.** The Y-axis reports the percentage of reads for the two parameters for each sample, grouped by the fixation protocol. P-value is calculated with paired T-test. **C OncoPrint of variants detected by the OCAV3 panel.** Gene names and relative frequency of mutations identified in the 9 CRCs are reported in the double y-axis. Top annotation show the number of variant/patient for each genes; whereas annotations at the bottom report the fixation type and the sample ID.

Amplicon-based libraries also allow the calculation of the Median Absolute Pairwise Difference, which consists of the difference between the quantity (as log_2_ of read count ratio) of adjacent amplicons, as a measure of “amplification noise”. In line with previous data, libraries from NBF tissues showed the highest level of noise, significantly higher than ADF and *cold*ADF (Supplementary Figure 2B, Supplementary Table 2). Similarly to previous data, libraries from GAF tissues showed a medium level noise.

Sequencing results are reported in the OncoPrint in Figure 2C. OCAV3 somatic variants are reported in Supplementary Table 3, together with the alteration identified in the TSO500 sequencing. The hepatoma did not harbor any variants in any of the parallel samples. When focusing on CRCs, we observed that CRC#9 showed more variants than the other tumor samples. At the same time, CRC#9 samples with different fixation harbored variability in terms of type and quantity detected variants. In addition, scattered and rare discrepancies were identified within parallel samples (different fixation protocols) of other CRCs (discussed below by integration of a cross-comparison with the hybrid-capture based data).

### Impact of the different fixations over a CGP based on a hybrid-capture DNA targeted sequencing

Comparative results for sequencing with the TruSight Oncology 500 panel were divided into library sequencing QCs and sequencing results.

In line with OCAv3, median depth was not influenced by the fixation type (Supplementary Table 4) and read length comparison confirmed longer reads for ADF and *cold*ADF, with NBF libraries being significantly shorter compared to other fixatives Supplementary Table 4).

The TSO500 is a Unique Molecular Identifiers (UMI)-based panel, in which these short random nucleotide sequences are added to each molecule of a sample prior to PCR, to reduce the impact of PCR-duplicates. By grouping the UMI deduplicated reads into families, the pipeline returns the UMI family size (larger size → lower PCR errors) and the percentage of chimeric reads (i.e., PCR duplicates). No differences were observed within the 4 fixatives for the percentage of chimeric reads, however ADF and *cold*ADF derived-libraries showed the highest UMI-family size, compared with GAF and NBF (Supplementary Table 4).

The coding size of TSO500 consists of 1.27 Mb in size, and we wondered whether different fixatives could influence the total region sequenced. In line with the lower quality of the libraries, NBF showed a significantly smaller region sequenced, with both ADF-based fixatives showing the best performance (Figure 3A and Supplementary Table 4). TSO500 also evaluates a total of 120 MS loci for the MS instability test. We observed a significantly lower number of evaluable MS loci in NBF, compared to all the other fixatives (Supplementary Table 4).

**Figure 3.**
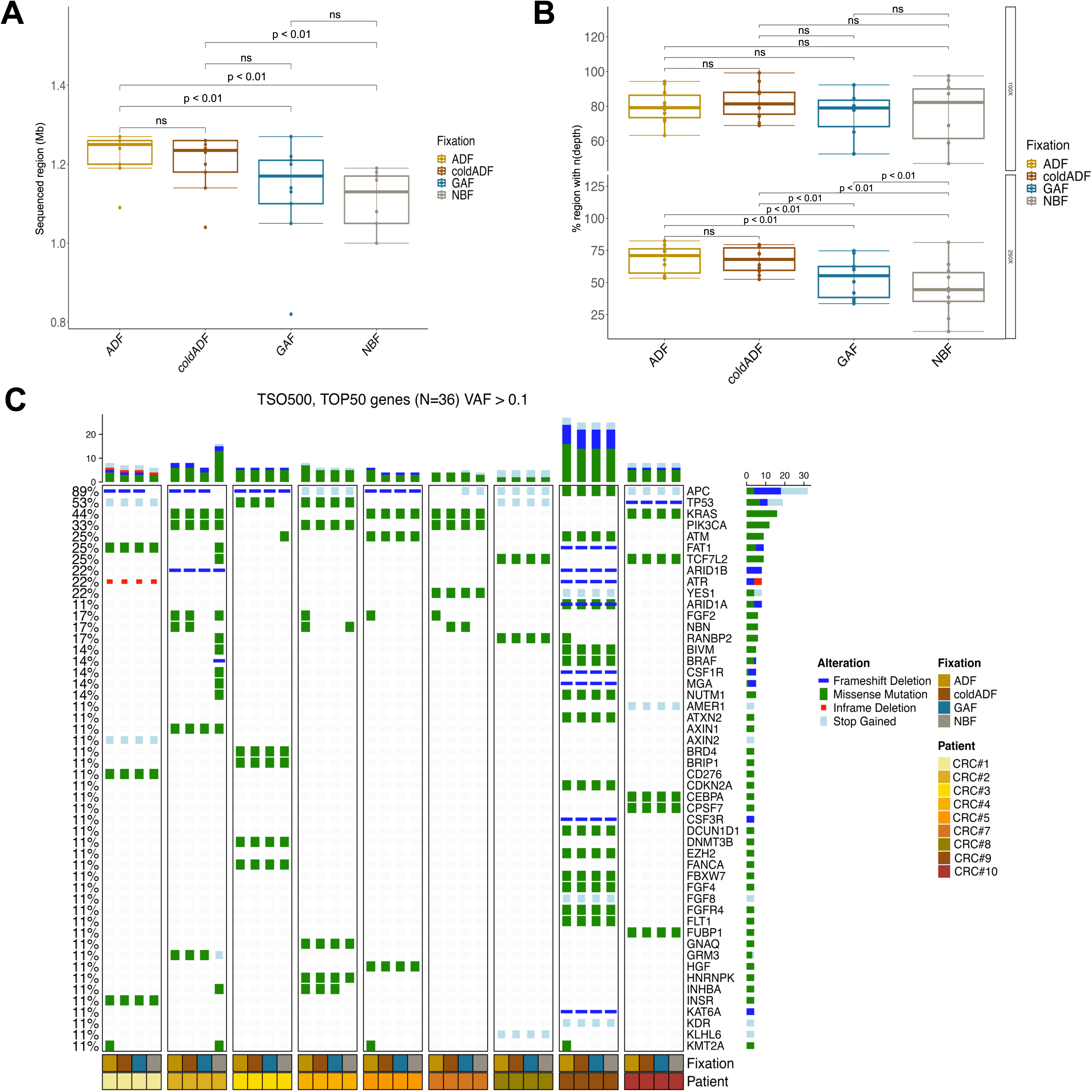
Output data by the TSO500 panel. A) Boxplot of the exonic sequenced regions. The Y-axis reports the mean Mb sequenced for each sample, grouped by the fixation protocol. P-value is calculated with paired T-test. **B) Boxplots of the TSO500 target region sequenced at 100X and 250X.** The Y-axis reported the percentage of target region covered at 100X and 200X for each sample, grouped by the fixation protocol. P-value is calculated with paired T-test. **C) OncoPrint of the variants detected in the 50 most mutated genes.** Gene names and relative frequency of mutations identified in the 9 CRCs are reported in the double y-axis. Top annotations show the number of variant/patient for each genes; whereas annotation at the bottom reported the fixation type and the sample ID.

The analysis of the reads on-target demonstrated a significantly higher quantity of off-targets in the libraries stemming from NBF fixed samples, thus affecting the uniformity of coverage at 250X that was significantly lower for NBF samples compared to the others (Figure 3B).

The OncoPrint in Figure 3C reports the 50 most frequently mutated genes with somatic variants harboring a VAF of at least 0.1. The hepatoma samples were all WT. In line with the OCAv3 results, CRC#9 showed a high mutational load (ADF=66.3, *cold*ADF =76.1, GAF=66.6, NBF=68.7) and a high level of MSI (ADF=52.9%, *cold*ADF =48.1%, GAF=65.7%, NBF=60.0%). In addition, visual inspection of the OncoPrint revealed an increased number of variants for CRC#2 NBF-fixed only, compared to the other CRC#2 samples.

Similar to OCAv3 panel results, we observed scattered differences across the samples. To systematically evaluate similarities/discrepancies we planned specific analyses. First, we performed a cross-comparisons of somatic variants affecting the genomic regions covered by OCAv3 and TSO500, to assess the gain or the loss of variants in the main cancer related genes. Second, based on TSO500 data we also evaluated intra-patient differences through the calculator of the Jaccard coefficient for each single base alteration (both germline and somatic with a VAF>=0.1, both synonymous and nonsynonymous). Third, we assessed the variant type and predicted the mutational signature profiles.

For the cross-comparison analysis, we excluded the CRC#9 due to the high number of variants and possible intrinsic heterogeneity of variants in MSI lesions.

We observed a substantial overlap with 17 unique variants shared by all the samples and confirmed by both panels, however five genes showed a different status across samples and/or across panels (Figure 4A). Of these discordant results, 4 involved samples purified from NBF-fixed tissues.

**Figure 4.**
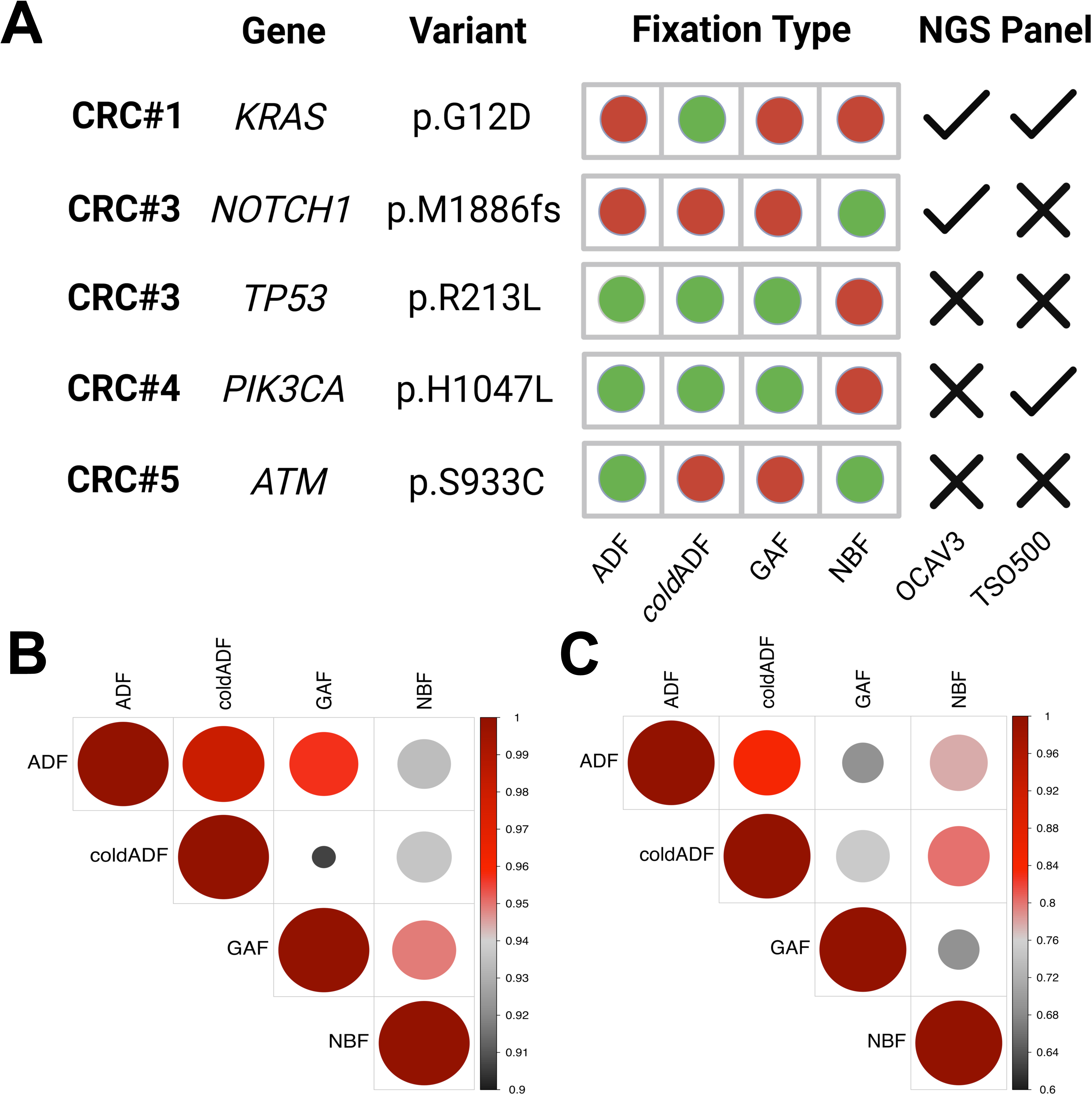
Correlations across samples and gene panels. A) Representation of the sequencing discrepancy within samples and panels. Each raw represents a patient with a sequencing incongruity. We report the affected gene, the gene variant, the fixation type and the NGS panel. Red bullets identify the discrepancy, tick and cross symbols define the panel in which the alteration was wither identify or missing, respectively. **B) Correlation plot of the JI germline variant comparison.** Size and color of the bullet**s** summarize the similarity within the fixation types. Red and larger dots are proper of fixatives with similar nucleotide changes, whereas grayish and smaller bullets represent fixatives with more variable changes. Scale bar of JI for germline comparison ranges from 0.9 and 1.0. **C) Correlation plot of the JI somatic variant comparison.** Size and color of the bullets summarize the similarity within the fixation types. Red and larger dots are proper of fixatives with similar nucleotide changes, whereas grayish and smaller bullets represent fixatives with more variable changes. Scale bar of JI for somatic comparison ranges from 0.6 and 1.0.

CRC#3-NBF harbored a *NOTCH1* SNV (p.M1886fs) by OCAv3 sequencing, not identified in the other CRC#3 samples and neither in the CRC#3-NBF sequenced with theTSO500. In the same sample the *TP53* p.R213L mutation shared by all the other lesions was not detected (either by OCAv3 or TSO500). The *PIK3CA* pH1047L was present in all CRC#4 samples by both panels, except for the OCAv3 analysis of the NBF-fixed sample.

In addition, an *ATM* variant (p.S933C) was detected only in 2/4 CRC#5 samples: both OCAv3 and TSO500 analyses did not identify this variant in the NBF and ADF fixed specimens.

Finally, CRC#1 *cold*ADF-fixed showed a private and sub-clonal *KRAS* p.G12D, not detected in the other samples.

TSO500 intra-panel comparison of synonymous and nonsynonymous variants allowed us to calculate the Jaccard index (JI) within the fixation types, a quantitative measurement of similarity across the fixatives. By considering both germline and somatic variants, all the fixation showed a high level of correlation (JI>0.90). Of note, ADF4 and *cold*ADF4 showed the closest similarity, whereas GAF and NBF-fixed samples showed a smaller concordance (Figure 4B). This trend is even clearer considering only somatic variants: GAF samples showed the lowest JI compared with all the other samples (0.75 with *cold*ADF and 0.69 for both ADF and NBF), with ADF retaining a strong correlation with *cold*ADF (0.84), and NBF (0.78), in line with the JI = 0.8 between *cold*ADF and NBF (Figure 4C).

To qualitatively evaluate genetic features within the different fixations, we compared the variant type (e.g SNVs, deletions and insertions) and the variant classification (e.g. missense, synonymous, frameshift, stop gained variant and in frame deletions). A significantly increased number of SNVs is present in the NBF fixed lesions, associated also with an increased number of missense, synonymous, frameshift, stop gained variants (p<0.01 comparted to all the other fixatives, Supplementary Figure 3A-B). No differences were identified across the other fixation protocols.

Finally, we predicted the mutational signature profiles for each fixation type. The 96-matrix profile showed similar but non completely overlapping substitutions (Figure 5A). By quantifying the relative contribution of each signature, we identified similar weight for signature 6 and 15 for each fixation type, related to the MSI phenotype. Signature 1, aging-related but also referred to FFPE artifacts showed a significant reduction from NBF (37%) to ADF (30%) and GAF (31%) fixation, reaching the 17% in the *cold*ADF samples (Figure 5B).

**Figure 5.**
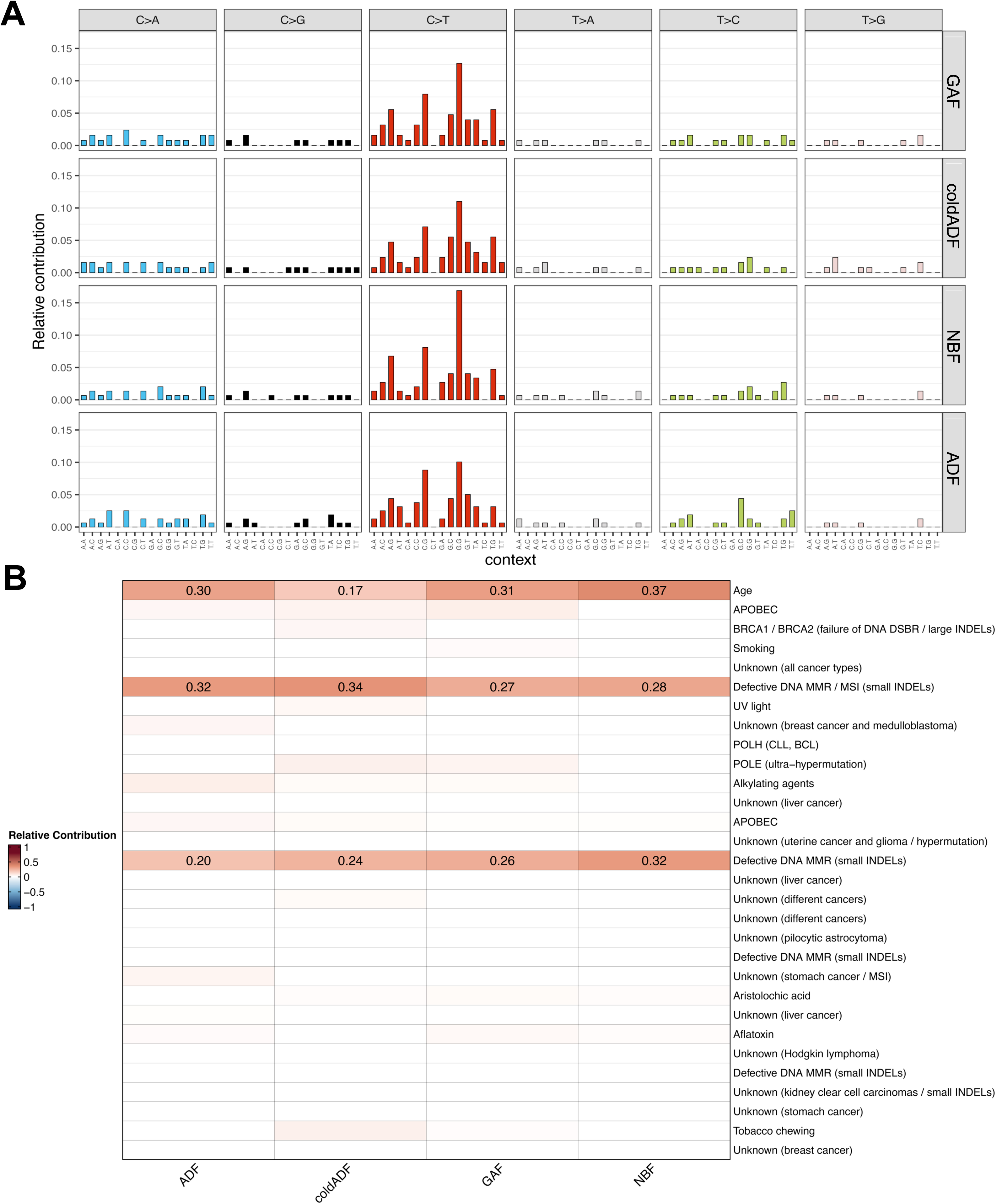
Mutational signatures derived from the samples. A) Representation of mutational signatures by using the 96-matrix profile, based on substitutions identified for the 4 fixation types. The picture reports all the raw substitutions for all samples, grouped for the fixation types. **B) Relative contribution of each predicted COSMIC V2 signatures for each group.** We estimated the mutational signature contribution, comprising all the variants with a VAF>0.05. We reporte the description of the signatures, sorted up-to-down from signature #1 to signature #30.

## DISCUSSION

Here we report a detailed analysis on the effect of different fixation protocols over DNA integrity/preservation of tissue samples, thus providing evidence of the impact these pre-analytical features may have when performing CGP data analysis.

Precision medicine in Oncology is currently demanding a huge effort in extracting the most valuable information from tissue samples ^22, 23^. This is particularly important for advanced stage cancer patients. To fulfill this task, pathologists and molecular biologists use a combination of *in situ* techniques (immunohistochemistry and *in situ* hybridization) and molecular profiling by *in vitro* nucleic acid-based assays. The latter are currently best approached by targeted NGS, with commercially available panels that can be variable in terms of size (number of genetic alterations/genes that can be analyzed). Molecular diagnostics typically employs panels with a limited number of cancer-associated genes (up to 50), and a reference range that can be variable across panels. This strategy helps deconvolute the complexity of an NGS analysis on degraded DNA/RNA by focusing on a handful of targets that are specifically needed at present for treatment decision making. Nevertheless, the number and complexity of genetic alterations to be investigated is rapidly growing (even with an agnostic-fashion)^24–27^. In this scenario panels limited in size may not be fit for purpose, especially when screening for molecular alterations that could be of interest for ongoing clinical trials^24, 26^. Therefore, in several instances archival FFPE tissue samples undergo a CGP analysis, which is likely to expand its applicability in the next future. The challenge of a CGP approach is appreciated at two distinct levels: i) the success rate of the sequencing; ii) the complexity of data interpretation. Both features largely depend on the quality of nucleic acids.

In this study, we provide evidence of a critical improvement in DNA preservation, as compared to the classical NBF fixation, when using alternative aldehyde fixatives, still acting by cross-linking and providing a similar structural and antigenic preservation. We tested both ADF (Acid-deprived Formalin), obtained by the removal of the formic acid residues present in the commercial Formaldehyde reagent via ion-exchange resins, and GAF (Glyoxal, Acid Free), obtained from acidic Glyoxal by the same procedure of resin treatment. Both reagents already provided comparable results with standard FFPE samples when performing ancillary analyses such as IHC and to offer an improvement in DNA preservation^9, 10^. Here we aimed to comprehensively evaluate the impact of different fixation protocols on the performance of data analysis from a CGP approach.

We acted at different levels, from pre-analytical features to output sequencing results. DNA yield was equal across different extractions, even when normalizing the total DNA by the size of the lesions. The present study of DNA integrity confirmed and extended our previous report^9^ and showed a lower degree of DNA fragmentation for *cold-*ADF and ADF samples, which were also the most represented samples in the class with lower fragmentation of the clustering analysis generated from the fragment size distribution data. Similar results were obtained when comparing GAF, an acid-deprived solution of Glyoxal, and a buffered solution of the acid commercial Glyoxal. These results suggest a clear increment in terms of DNA purity and structural integrity when fixing tissues with acid deprivation. Of note, the highest level of purity and integrity was obtained with ADF and *cold*ADF, but tissues fixed in acid-deprived Glyoxal (GAF) were still superior to NBF as far as DNA purity and integrity is concerned.

The data are overall highlighting the crucial role played by buffered acids when present in fixatives and support the hypothesis, that can be summarized as “hidden acid theory”, where within the micro-environment represented by the cell nucleus phosphate radicals of nuclei acids can dislodge Sodium ions so that formic and other acidic residues are free to affect the DNA integrity^9^.

We also tested the effect of a fixation at low temperature (4°C) over fixation at room temperature. Confirming a previous study by our group^13^, we observed that the temperature degree is among the multiple factors bound to impact on DNA integrity.

We then moved to sequencing performance and data. To objectively assess potential differences across the distinct conditions here applied, we meticulously analyzed several parameters important for library preparation QCs, sequencing QCs and sequencing results. Overall, significantly longer reads were obtained in libraries from *cold*ADF samples with both the OCAv3 panel and the TSO500 panel. ADF-, GAF-and NBF-samples followed with the shortest reads demonstrated in NBF samples.

By analyzing the Oncomine Comprehensive v3 panel data, ADF and *cold*ADF derived libraries showed the lowest level of noise and the highest levels of uniformity, followed by GAF and NBF fixed tissues. The TSO500 panel data confirmed the data uniformity. In addition, with this panel we observed the best performance in terms of total region sequenced for ADF and *cold*ADF samples, whereas NBF samples showed a significantly smaller region sequenced. Along the same lines, NBF samples displayed a significantly lower number of evaluable MS loci compared to all the other fixatives.

Despite a formally successful sequencing in all the samples, these QC and sequencing data analyses suggest an increment in terms of quality of library generation and sequencing outputs from NBF– to GAF– to ADF– and *cold*ADF-fixed samples.

One may wonder whether these features affect data reporting and interpretation.

Most of the genetic alterations identified were shared by all the samples in each tumoral lesion. However, five genes showed a different status across samples and/or across panels. An important observation is that 4 of these discordant results involved samples purified from NBF-fixed tissues, thus suggesting a possible artefactual origin. Although the tissues here analyzed were sampled in parallel from the same region of a given tumoral lesion and we may assume that heterogeneity is highly unlikely, we cannot exclude this alternative explanation for such discordances. We favor arteficts over biological heterogeneity based on additional data derived from the TSO500 panel. A significantly increased number of SNVs was present in the NBF fixed lesions, whereas no differences were identified across the other fixation protocols. Finally, when mutational signatures were predicted Signature 1 (aging-related but also referred to FFPE artifacts^28^) showed the highest values in NBF samples (37%) and the lowest values in *cold*ADF samples (17%).

In conclusion, fixation is the preliminary, yet central, process in histological processing leading to the production of paraffin-embedded tissue blocks, the ultimate product constituting the backbone of pathology archives. Different modalities of tissue fixation lead to differential degrees of DNA integrity, which impacts on the output of comprehensive genomic profiling. Acid-deprived fixatives guarantee the highest DNA preservation overall, thus suggesting the possible implementation of even more complex molecular profiling on tissue samples.

## Data Availability

All data produced in the present study are available upon reasonable request to the authors

## Acknowledgements

This research was funded by FONDAZIONE AIRC under IG 2019 – ID. 22850 project – P.I. Marchiò Caterina. Part of the analyses were supported by Alleanza Contro il Cancro (Alliance Against Cancer), Ricerca Corrente 2021 to Working Group Pathology and Biobanking and “Progetto ACCORD, il Registro ACC delle *Omiche*”.

We also acknowledge partial funding by FONDAZIONE AIRC (Associazione Italiana per la Ricerca sul Cancro) under 5 per Mille 2018—ID 21091 program—Group Leader: CM and FPRC 5xmille 2018 Ministero Salute, project “ADVANCE/A-Bi-C”: Italian Ministry of Health, Ricerca Corrente 2021 to MC.

We would like to thank the technical and medical staff of the Pathology Unit of Candiolo Cancer Institute for support in tissue sample processing and manipulation for molecular downstream analyses.

## SUPPLEMENTARY FIGURE LEGENDS

**Supplementary Figure 1.**
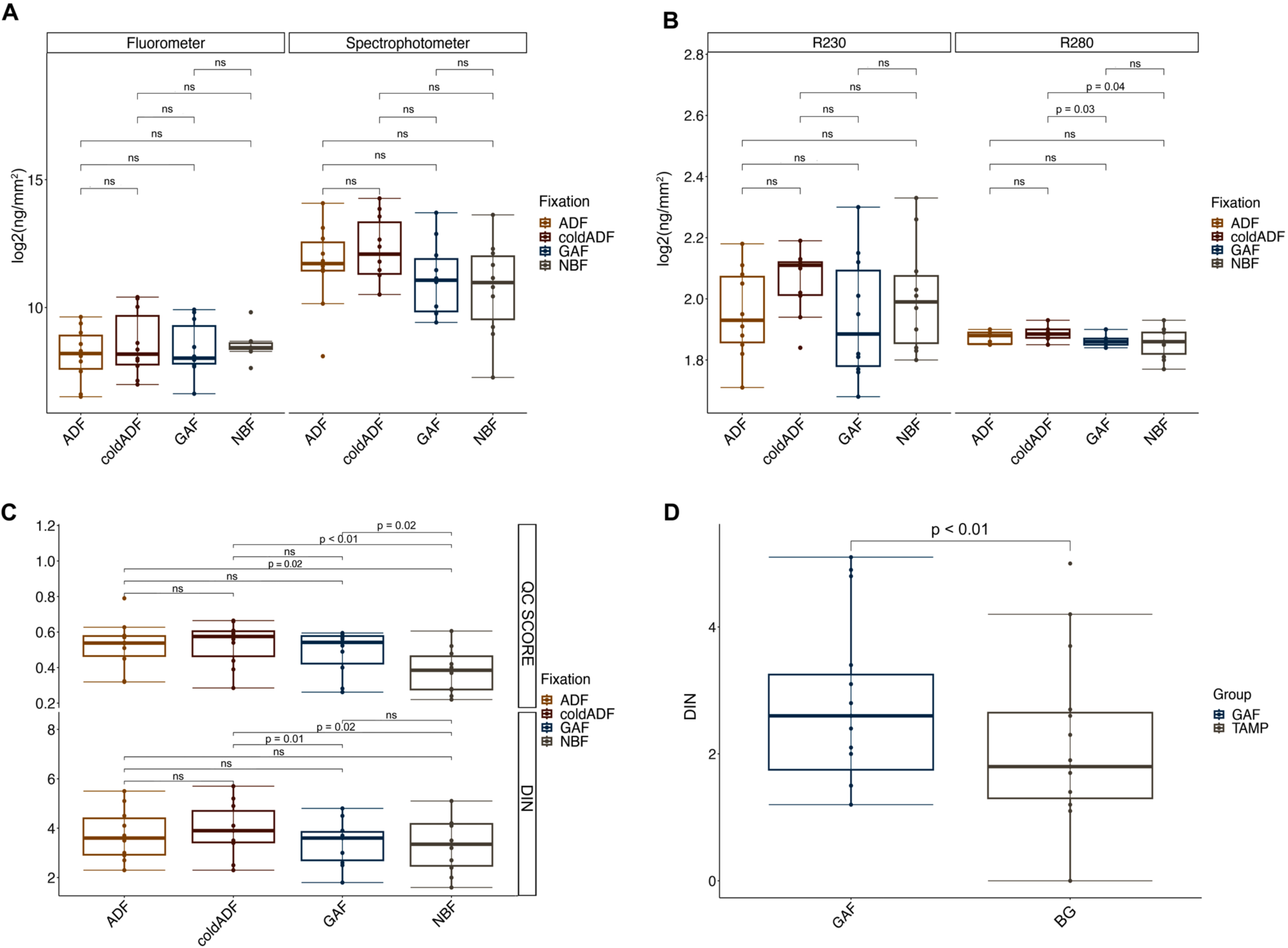
A) Boxplot of the DNA extraction yield. The Y-axis reports the _log2_(ng/mm^2^) for each sample, grouped by the fixation protocol, for both spectrophotometer and fluorometer. P-value is calculated with paired T-test. B) Boxplot of the DNA extraction absorbance ratios. The Y-axis reports the R280 and R230 absorbance values for each sample, grouped by the fixation protocol. P-value is calculated with paired T-test. C) Boxplot of the DNA quality controls. The Y-axis reports the DIN and QC Score values for each sample, grouped by the fixation protocol. P-value is calculated with paired T-test. D) Boxplot of the DIN comparison for GAF versus Buffered Gyloxal (BG).

**Supplementary Figure 2.**
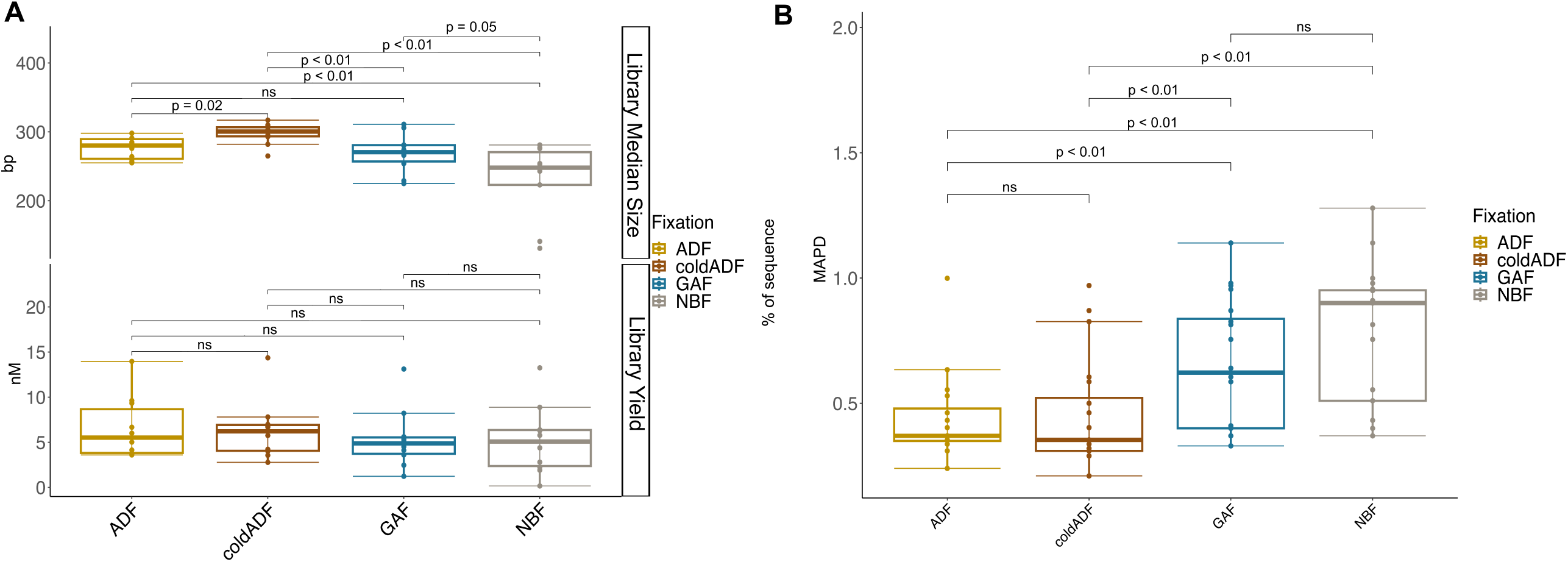
A) Boxplot of OCAV3 library median size and library yield. B) Boxplot of OCAV3 MAPD.

**Supplementary Figure 3.**
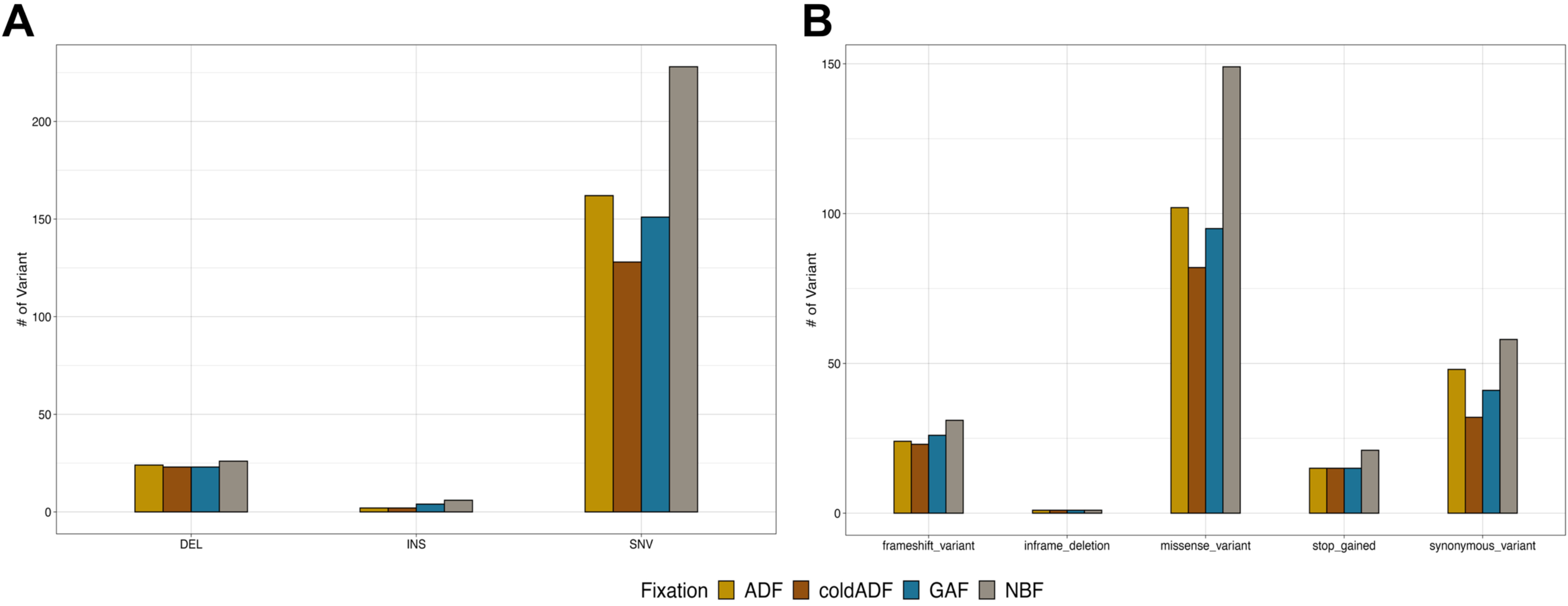
A). Comparison of Variant Classification (VC) distribution among the fixation cohorts; B) Comparison of Variant Type (VT) distribution among the fixation cohorts.

## REFERENCES

1 Mathieson W and Thomas GA. Why Formalin-fixed, Paraffin-embedded Biospecimens Must Be Used in Genomic Medicine: An Evidence-based Review and Conclusion. J Histochem Cytochem 68, 543–552, doi:10.1369/0022155420945050 (2020).

2 Cucco F, Clipson A, Kennedy H, Sneath Thompson J, Wang M, Barrans S et al. Mutation screening using formalin-fixed paraffin-embedded tissues: a stratified approach according to DNA quality. Lab Invest 98, 1084–1092, doi:10.1038/s41374-018-0066-z (2018).

3 Kim S, Park C, Ji Y, Kim DG, Bae H, van Vrancken M et al. Deamination Effects in Formalin-Fixed, Paraffin-Embedded Tissue Samples in the Era of Precision Medicine. J Mol Diagn 19, 137–146, doi:10.1016/j.jmoldx.2016.09.006 (2017).

4 Sah S, Chen L, Houghton J, Kemppainen J, Marko AC, Zeigler R et al. Functional DNA quantification guides accurate next-generation sequencing mutation detection in formalin-fixed, paraffin-embedded tumor biopsies. Genome Med 5, 77, doi:10.1186/gm481 (2013).

5 Hedegaard J, Thorsen K, Lund MK, Hein AM, Hamilton-Dutoit SJ, Vang S et al. Next-generation sequencing of RNA and DNA isolated from paired fresh-frozen and formalin-fixed paraffin-embedded samples of human cancer and normal tissue. PLoS One 9, e98187, doi:10.1371/journal.pone.0098187 (2014).

6 Amemiya K, Hirotsu Y, Oyama T, and Omata M. Relationship between formalin reagent and success rate of targeted sequencing analysis using formalin fixed paraffin embedded tissues. Clin Chim Acta 488, 129–134, doi:10.1016/j.cca.2018.11.002 (2019).

7 Endrullat C, Glokler J, Franke P, and Frohme M. Standardization and quality management in next-generation sequencing. Appl Transl Genom 10, 2–9, doi:10.1016/j.atg.2016.06.001 (2016).

8 Kaneko Y, Kuramochi H, Nakajima G, Inoue Y, and Yamamoto M. Degraded DNA may induce discordance of KRAS status between primary colorectal cancer and corresponding liver metastases. Int J Clin Oncol 19, 113–120, doi:10.1007/s10147-012-0507-4 (2014).

9 Berrino E, Annaratone L, Detillo P, Grassini D, Bragoni A, Sapino A et al. Tissue Fixation with a Formic Acid-Deprived Formalin Better Preserves DNA Integrity over Time. Pathobiology, 1–11, doi:10.1159/000525523 (2022).

10 Bussolati G, Annaratone L, Berrino E, Miglio U, Panero M, Cupo M et al. Acid-free glyoxal as a substitute of formalin for structural and molecular preservation in tissue samples. PLoS One 12, e0182965, doi:10.1371/journal.pone.0182965 (2017).

11 Annaratone L, Marchio C, and Sapino A. Tissues under-vacuum to overcome suboptimal preservation. N Biotechnol 52, 104–109, doi:10.1016/j.nbt.2019.05.007 (2019).

12 Bussolati G, Chiusa L, Cimino A, and D’Armento G. Tissue transfer to pathology labs: under vacuum is the safe alternative to formalin. Virchows Arch 452, 229–231, doi:10.1007/s00428-007-0529-x (2008).

13 Berrino E, Annaratone L, Miglio U, Maldi E, Piccinelli C, Peano E et al. Cold Formalin Fixation Guarantees DNA Integrity in Formalin Fixed Paraffin Embedded Tissues: Premises for a Better Quality of Diagnostic and Experimental Pathology With a Specific Impact on Breast Cancer. Front Oncol 10, 173, doi:10.3389/fonc.2020.00173 (2020).

14 Ryska A. SA, Landolfi S., Sansano Valero I., Ramony Cajal S., Pedro Oliveira. Detillo P., Lianas L., Frexia F., Nicolosi P.A., Monti T., Bussolati B., Marchiò C., Bussolati G. Glyoxal Acid-Free (GAF) histological fixative is a suitable alternative to formalin – results from an open label comparative non-inferiority study. medRxiv Preprint, doi: https://doi.org/10.1101/2023.05.24.23290451 (2023).

15 Sartore-Bianchi A, Pietrantonio F, Lonardi S, Mussolin B, Rua F, Crisafulli G et al. Circulating tumor DNA to guide rechallenge with panitumumab in metastatic colorectal cancer: the phase 2 CHRONOS trial. Nat Med 28, 1612–1618, doi:10.1038/s41591-022-01886-0 (2022).

16 Berrino E, Annaratone L, Bellomo SE, Ferrero G, Gagliardi A, Bragoni A et al. Integrative genomic and transcriptomic analyses illuminate the ontology of HER2-low breast carcinomas. Genome Med 14, 98, doi:10.1186/s13073-022-01104-z (2022).

17 Berrino E, Aquilano MC, Valtorta E, Amodio V, Germano G, Gusmini M et al. Unique Patterns of Heterogeneous Mismatch Repair Protein Expression in Colorectal Cancer Unveil Different Degrees of Tumor Mutational Burden and Distinct Tumor Microenvironment Features. Mod Pathol 36, 100012, doi:10.1016/j.modpat.2022.100012 (2023).

18 Berrino E, Filippi R, Visintin C, Peirone S, Fenocchio E, Farinea G et al. Collision of germline POLE and PMS2 variants in a young patient treated with immune checkpoint inhibitors. NPJ Precis Oncol 6, 15, doi:10.1038/s41698-022-00258-8 (2022).

19 Diaz-Gay M, Vila-Casadesus M, Franch-Exposito S, Hernandez-Illan E, Lozano JJ, and Castellvi-Bel S. Mutational Signatures in Cancer (MuSiCa): a web application to implement mutational signatures analysis in cancer samples. BMC Bioinformatics 19, 224, doi:10.1186/s12859-018-2234-y (2018).

20 Cereda M, Gambardella G, Benedetti L, Iannelli F, Patel D, Basso G et al. Patients with genetically heterogeneous synchronous colorectal cancer carry rare damaging germline mutations in immune-related genes. Nat Commun 7, 12072, doi:10.1038/ncomms12072 (2016).

21 Alexandrov LB, Nik-Zainal S, Wedge DC, Campbell PJ, and Stratton MR. Deciphering signatures of mutational processes operative in human cancer. Cell Rep 3, 246–259, doi:10.1016/j.celrep.2012.12.008 (2013).

22 Annaratone L, De Palma G, Bonizzi G, Sapino A, Botti G, Berrino E et al. Basic principles of biobanking: from biological samples to precision medicine for patients. Virchows Arch 479, 233–246, doi:10.1007/s00428-021-03151-0 (2021).

23 Force APT. Pathology: Hub and Integrator of Modern, Multidisciplinary [Precision] Oncology. Clin Cancer Res 28, 265–270, doi:10.1158/1078-0432.CCR-21-1206 (2022).

24 Fountzilas E, Tsimberidou AM, Vo HH, and Kurzrock R. Clinical trial design in the era of precision medicine. Genome Med 14, 101, doi:10.1186/s13073-022-01102-1 (2022).

25 Gouda MA, Nelson BE, Buschhorn L, Wahida A, and Subbiah V. Tumor-Agnostic Precision Medicine from the AACR GENIE Database: Clinical implications. Clin Cancer Res, doi:10.1158/1078-0432.CCR-23-0090 (2023).

26 Perez EA. In: Ramirez AG and Trapido EJ editors. Advancing the Science of Cancer in Latinos Cham (CH): 2020. p. 113–123.

27 Yan L and Zhang W. Precision medicine becomes reality-tumor type-agnostic therapy. Cancer Commun (Lond) 38, 6, doi:10.1186/s40880-018-0274-3 (2018).

28 Guo Q, Lakatos E, Bakir IA, Curtius K, Graham TA, and Mustonen V. The mutational signatures of formalin fixation on the human genome. Nat Commun 13, 4487, doi:10.1038/s41467-022-32041-5 (2022).

